# Longitudinal association between depressive symptoms and self-directed passive aggression: A random intercept cross-lagged panel analysis

**DOI:** 10.1101/2022.04.12.22273775

**Authors:** Christian G. Schanz, Monika Equit, Sarah K. Schäfer, Tanja Michael

## Abstract

**Background:** Self-directed passive aggression (SD-PAB) is defined as any behaviour harming one-self by inactivity and omission of own needs. Depressive disorders are a severe mental disorder that results from the interaction between stress exposure, coping strategies, and vulnerability. Previous cross-sectional studies found SD-PAB to be associated with depressive symptoms and to represent a mediator of the relationship between cognitive risk factors and depressive symptoms. Therefore, SD-PAB may be a potential target of prevention or treatment in the context of depressive disorders. However, prospective studies on the relationship between depressive symptoms and SD-PAB are lacking. The current study aimed at closing this gap by examining the associations of subjective stress, SD-PAB, and depressive symptoms cross-sectionally and over time.

**Method:** In two assessment cohorts students participated three times [M1: start of the semester (*n* = 352); M2: start of the exam period (*n* = 293); M3 = end of the exam period (*n* = 276)] in an online survey (depressive symptoms; self-perceived stress; SD-PAB). Cross-sectional data was analysed using regression models. Longitudinal data was analysed using Random Intercept Cross-lagged Panel Models.

**Results:** Across all time points, SD-PAB demonstrated a unique cross-sectional association with depressive symptoms when controlled for self-perceived stress (*β* = .27 – .33; all *p*’s < .001). Furthermore, at M2 [*β* = .14, *t*(289) = 3.71, *p* < .001] and M3 [*β* = .15, *t*(272) = 3.51, *p* < .001] the relationship between depressive symptoms and self-perceived stress was stronger for individuals reporting higher levels of SD-PAB. Depressive symptoms at M1 are a marginal significant predictor of SD-PAB at M2 (*β* = .31; *p* = .067) and depressive symptoms at M2 are a marginal significant predictor for SD-PAB at M3 (*β* = .17; *p* = .074). However, there was no evidence for SD-PAB predicting the course of depressive symptoms.

**Conclusion:** SD-PAB may represent a symptom of depressive disorders and a moderator of unsuccessful stress coping but does not predict the course of depressive symptoms over time.

## Introduction

Self-directed aggressive behaviour (SD-AB) is defined as any behaviour intending to harm oneself directly or indirectly through active engagement (active SD-AB) or by omission [passive SD-AB; (1–3)]. SD-AB may serve different purposes in the short-term [e.g. affect regulation; (4)], but is normally prevented by mental barriers serving to maintain mental and physical integrity of oneself [e.g. pain barrier, positive self-barrier; (5)]. SD-AB is particularly prevalent in individuals with high levels of stress (6), increased impulsivity (7,8), and reduced self-compassion (9,10). Individuals with depressive disorder (DD) are at high risk for SD-AB (11– 13). Several studies show an association between DD and active SD-AB (14). Furthermore, two recent studies demonstrate a moderate to strong association between passive SD-AB and depressive symptoms in inpatients, outpatient as well as student samples (15,16).

DD is characterized by depressed mood and loss of interest or pleasure (17). Among all mental disorders, DD accounts for most disability-adjusted life years (DALYs) (18) and is associated with huge economic costs (19). Across the lifespan the prevalence of DD is high (10.6% - 19.8%), with first episodes most frequently occurring in early adulthood (20). For example, college students experiencing high levels of academic stress have an increased risk for the onset of DD (21–25). According to cognitive behavioural theories based on diathesis-stress models, DD is caused inter alias by loss of reinforcement (26,27), dysfunctional attitudes (28– 31), and negative attributional styles (32–34). These factors also play a key role in the self-control model of depression (35), which postulates reduced self-reward and increased self-punishment as key components of the development of DD.

Based on Kanfer’s (36) self-control theory, the self-control model of depression supposes that decreased self-reinforcement and increased self-punishment derive from dysfunctional self-monitoring and self-evaluation processes [i.e. due to dysfunctional attitudes and negative attributional style;(35)]. Self-punishment reflects active SD-AB and is seen in patients with DD in form of self-blame, non-suicidal self-harm, and suicidal behaviour (37,38). Reduction of self-reinforcement is a form of passive SD-AB and contributes to the core symptoms of DD (depressed mood and loss of interest or pleasure) by the loss of reinforcement (26,27). Therefore, active and passive SD-AB could constitute risk factors for the development of DD. However, one may also assume different associations between SD-AB and DD: Some studies revealed active SD-AB as predictor for DD (39,40), others found DD to be a predictor for active SD-AB (41–44), or DD and active SD-AB to interact (45). However, other studies found cross-sectional but not longitudinal associations between active SD-AB and DD (46–48). In sum, there is tentative evidence that DD predicts active SD-AB, which, if confirmed, would indicate the need of prevention strategies for active SD-AB in patients suffering from DD. To the best of our knowledge, to date, no prospective study has investigated the relationship between passive SD-AB and DD. Because insight in the longitudinal association of passive SD-AB and DD could be relevant for prevention and treatment of both passive SD-AB and DD, the current prospective study examined their association over the course of one semester in a student sample.

## Study aims

The current study has three aims: First, to replicate previous findings (15,16) on the association between passive SD-AB and depressive symptoms. Second, to analyse if passive SD-AB moderates the association between subjective stress and depressive symptoms, which would support the notion of SD-AB being a dysfunctional self-regulation strategy. Third, to examine if baseline SD-AB and depressive symptoms severity predict changes in one-another in the course of a moderate stressful event (an exam phase). Thereby, the current study will help to clarify the question whether passive SD-AB is a risk-factor for DD or a consequence of DD symptoms.

## Methods

### Procedure and participants

Participants were recruited in two waves (Preregistration-Number: DRKS00014610 and DRKS00015008). Wave 1 was conducted in the autumn term 2018 and Wave 2 in the autumn term 2019 at the Saarland University. For both waves, adult first semester students (age ≥ 18 years) were recruited. Wave 1 used a sample of psychology students, while Wave 2 included undergraduate students of psychology, biology, and educational science. Recruitment took place in first semester lectures. Subjects received course credits or monetary allowance (€ 15) for their participation. See Table 1 for sample characteristics.

**Table 1.** Sample characteristics

|  | M1 | M2 | M3 |
| --- | --- | --- | --- |
| N | 352 | 293 | 276 |
| Age (SD) | 20.92 (3.39) | 21.04 (3.64) | 21.03 (4.16) |
| % female | 79.3 | 79.5 | 79.7 |
| BDI-II | 11.29 (7.30) | 14.00 (9.01) | 12.21 (8.83) |
| GSI | 0.74 (0.52) | 0.78 (0.58) | 0.73 (0.58) |
| TICS-SF | 34.72 (8.43) | 37.60 (9.66) | 36.79 (9.76) |
| TPA-SD | 2.51 (0.64) | 2.41 (0.65) | 2.43 (0.70) |

*Note.* BDI-II = Beck Depression Inventory – II; GSI = Global Severity Index of the Brief Symptom Checklist; TICS-SF = Trier Inventory of Chronic Stress – Short Form; TPA-SD = Test of Passive Aggression – self directed; M1 – M3 = Measure point 1 – 3.

Assessment periods of both waves comprised three time points: At the start of the semester (measure point 1, M1), two weeks before the first exam of the semester (measure point 2, M2), and immediately after the final exam of the semester (measure point 3, M3). Participants received an e-mail as reminder for study participation and completed the online survey. Due to the rise of COVID-19 in Germany all pending exams at the Saarland University were cancelled in March 2020. Therefore, all participants which had not completed their last exam at that date (57.51%) were invited for M3 of Wave 2 at the day of exam cancellation.

For all assessments, the online survey platform SoSci Survey (49) was used for data collection. After giving written informed consent in accordance with the Declaration of Helsinki (50), the participants completed the short form of the the Trier Inventory for Chronic Stress [TICS-SF 4/12/2022 10:59:00 AM], the Beck Depression Inventory II [BDI-II (51)], the Brief Symptom Checklist [BSCL (52)], and the Test of Passive Aggression [TPA (15)].

### Measurements

#### Stress levels

Perceived subjective stress levels of the last month were assessed using the short form of the Trier Inventory for Chronic Stress [TICS (53)]. The TICS is a valid and reliable measure for perceived stress (54). The TICS-SF comprises 12 items selected from the one factor solution of the long form. Internal consistencies of the TICS-S were excellent across all assessments (*α* = .91 – .91).

#### Depressive symptoms

Depressive symptoms were assessed with the BDI-II (51). The 21 items of the BDI-II represent depressive symptoms based on the fourth version of the Diagnostic and Statistical Manual of Mental Disorders [DSM IV (55)]. The BDI-II is a valid and reliable measure of depressive symptoms (56), with very good internal consistencies in the current study (*α* = .83 – .89)

#### Psychopathological symptoms

General symptom severity was assessed using the BSCL(52). The BSCL consists of 53 items and its Global Severity Index (GSI) is a reliable and valid measure for general symptom intensity (57,58). The internal consistency of GSI was excellent across all assessments (*α* = .95 – .96).

#### Self-directed passive-aggressive behaviour

Passive SD-AB was measured using the TPA (15). Its passive SD-AB scale (TPA-SD) is a valid and reliable 12-item scale (59), which demonstrated acceptable to good internal consistencies in the current study (*α* = .77 – .84).

### Data Analysis

Regression analyses, *t*-tests for dependent samples and *Pearson Correlations* were conducted using IBM SPSS Statistics 25 (60). Moderator and mediator analyses were performed using *Process Macro* [95% confidence interval, number of bootstrapping samples = 5000; (61)]. Cross lagged panel models (CLPM) and Random Intercept CLPM (RI-CLPM) were estimated in *R* using the *riclpmr* package (62). CLPM have been frequently used for analysing data on the longitudinal course of the relationship between active SD-AB and DD (41,44). However, CLPM have been criticised recently for neglecting the trait-like stability of individual differences. RI-CLPM (62) are supposed to solve this problem by the inclusion of stable between-person differences. Fit of CLPM and RI-CLPM was compared using the *lavaan* package (63).

## Results

### Change of self-directed passive-aggressive behaviour, symptom severity and stress

Depressive symptoms increased from M1 to M2 [*t*(293) = 6.17, *p* < .001; *d* = 0.35] and decreased from M2 to M3 [*t*(276) = -4.14, *p* < .001; *d* = 0.18]. However, symptoms were more severe at M3 compared to M1 [*t*(267) = 2.40, *p* = .017; *d* = 0.15]. Similarly, general psychopathological symptoms increased from M1 to M2 [*t*(292) = 0.49, *p* <.001, *d* = 0.23) and decreased from M2 to M3 [*t*(267) = 5.48, *p* <.001; *d* = 0.23]. However, there was no difference between M1 and M3 [*t*(275) = 0.53, *p* = .600; *d* = 0.03]. Also perceived stress levels increased from M1 to M2 [*t*(292) = 6.80, *p* < .001, *d* = 0.34] and remained stable from M2 to M3 [*t*(266) = 1.64, *p* = .102; *d* = 0.08]. However, stress levels were significantly different between M1 and M3 [*t*(275) = 5.05, *p* < .001; *d* = 0.28]. TAP-SD decreased slightly between M1 and M2 [*t*(292) = -2.98; *p* = .003; *d* = -0.14] and M3 [*t*(275) = -2.05; *p* = .041; *d* = -.11], with no difference emerging between M2 and M3 [*t*(266) = 0.59; *p* = .555; *d* = 0.02].

### Associations between self-directed passive-aggressive behaviour, psychopathological symptom severity and perceived stress

Passive SD-AB was medium to strongly associated with depressive symptoms (*r* = .44 – .51, *p* <.001), general psychological symptoms (*r* = .41 – .49, *p* < .001), and perceived stress levels (*r* = .39 – .46, *p* < .001) across all assessments. Furthermore, even when controlled for general psychopathological symptoms, depressive symptoms explained an incremental amount of variance in passive SD-AB at all assessments (*β* = .27 – .33; *p* < .001).

### Moderation effect of self-directed passive-aggressive behaviour

For M2 [*β* = .14, *t*(289) = 3.71, *p* < .001] and M3 [*β* = .15, *t*(272) = 3.51, *p* < .001], but not M1 [*β* = .03, *t*(348) = 0.69, *p* = .49]), passive SD-AB moderated the association between perceived stress and depressive symptoms. For M2 and M3 more severe passive SD-AB were associated with a stronger relationship between perceived stress and depressive symptoms. Across all assessments, perceived stress and passive SD-AB explained each an incremental amount of variance of depressive symptoms, when controlled for each other and the moderator effect (*p* ≤ .003).

### Longitudinal association between self-directed passive-aggressive behaviour and depressive symptoms

RI-CLPM showed superior fit [*χ*^*2*^(1) = 0.12; *p* = .735] compared to the CLPM [*Δχ*^*2*^(3) = 21.20; *p* < .001]. In RI-CLPM, depressive symptoms at M2 were not predicted by passive SD-AB at M1 (*β* = .20; *p* = .150) and depressive symptoms at M3 were not predicted by passive SD-AB at M2 (*β* = .00; *p* = .972). However, passive SD-AB at M2 was predicted marginally by depressive symptoms at M1 (*β* = .31; *p* = .067) while passive SD-AB at M3 was predicted marginally by depressive symptoms at M2 (*β* = 17; *p* = .074).

## Discussion

The current study replicated previous findings on a moderate to strong association between passive SD-AB and depressive symptoms. Furthermore, passive SD-AB moderated the association between depressive symptoms and perceived stress in the course of a moderate stressor (i.e. exam phase). The RI-CLPM, which showed superior fit compared to the CLPM, identified depressive symptoms as a marginally significant predictor for passive SD-AB, while no evidence was found for the inverse relationship.

### Exam phase and depressive symptoms

In line with previous studies perceived stress was associated with depressive symptoms at all assessments (22–25). Both, depressive symptoms and perceived stress increased significantly from the start of the semester (M1) to the beginning of the exam phase (M2). Also, the decrease of depressive symptoms at the end of the exam phase is in line with previous finding demonstrating depressive symptoms to occur especially at the beginning of the exam phase(25). Therefore, as expected the beginning of the exam phase (M2) reflected a moderate stressor associated with an increase in depressive symptoms.

### Cross-sectional association between self-directed passive-aggressive behaviour and depressive symptoms

The results of our cross-sectional analyses correspond with our hypotheses, based on the self-control model of depression (35). In line with previous studies (16,59), passive SD-AB was moderate to strongly associated with depressive symptoms, even when general symptom severity was controlled for. Passive SD-AB also accounted for a significant amount of variance in depressive symptoms when controlled for perceived stress. Moreover, individuals with higher levels of passive SD-AB reacted with more severe depressive symptoms to stressful events. Thus, our results suggest that passive SD-AB is associated with ineffective stress management.

### Longitudinal relationship between self-directed passive-aggressive behaviour and depressive symptoms

The current longitudinal study is the first investigation using state-of-the-art methods for the analysis of prospective data examining whether passive SD-AB and depressive symptoms represent risk factors for each other. The RI-CLPM identified depressive symptoms as a marginally significant predictor of passive SD-AB, but not vice versa. This finding is in line with previous results, indicating depressive symptoms as predictor for active SD-AB (41–44). Therefore, passive SD-AB could be a relevant treatment target in DD.

## Limitations

Several limitations of the current study have to be considered. We investigated all associations in a student sample, which may have impeded the generalisability of our results, e.g. gender and age are distributed in a non-representative way. Moreover, the variance of clinical measures could have been restricted. However, previous studies identified female gender (64) and younger age (65) to constitute risk factors for SD-AB and student populations to be at high risk for the onset of DD (66). These findings demonstrate the relevance of SD-AB and DD research in those populations. Furthermore, depressive symptoms at M2 (but not at M1 and M3) matched the clinical cut-off for mild depression [BDI-II = 14 (51)], with BDI-II scores ranging from 1 to 51, thereby contradicting the assumption of severely restricted variances. Additionally, results regarding the association between passive SD-AB and depressive symptoms are in line with our previous studies in clinical samples with mean ages of *M* = 38.61 and *M* = 53.21 (15,59), demonstrating the comparability of results from student and patient samples. Depressive symptoms were assessed using the BDI-II, a self-report measure for severity of depressive symptoms. The BDI-II is a well validated and widely used measure of depressive symptoms with good sensitivity and specificity (56,67), however it is a screening tool and does not allow for categorical diagnostics of DD. Thus, the current study design allows for the examination of the course of depressive symptoms, but not for the onset of depressive episodes. Therefore, future studies should additionally use structured clinical interviews to enable the reliable and valid diagnosis of DD. Another possible limitation is the relatively small amount of perceived stress induced by the exam phase (M2 and M3). Although depressive symptoms and stress levels increased significantly from M1 to M2, effect sizes were small. Furthermore, in spite of stable stress across the period from M2 to M3, depressive symptoms declined in this period, indicating a short-term effect of the exam phase on depressive symptoms. Therefore, future studies should examine the longitudinal course of passive SD-AB, DD, and stress in high risk populations experiencing more severe stressors (e.g. police officers or firefighters).

## Conclusion

Cross-sectional analyses demonstrated that the relationship between depressive symptoms and subjective stress is stronger for individuals reporting higher passive SD-AB. Longitudinal analyses identified depressive symptoms as a predictor for the development of passive SD-AB levels. Therefore, individuals with mild levels of depression are at risk for passive SD-AB. Future studies should investigate a potential vicious cycle between passive SD-AB, dysfunctional coping, and depressive symptoms. Such studies may allow for the development of preventive interventions tailored to the specific needs of populations with mild levels of depression and high levels of perceived stress.

## Data Availability

All relevant data are within the manuscript and its Supporting Information files.

## Acknowledgments

Not applicable

## Supporting information captions

Not applicable

